# Robotic middle ear access for cochlear implantation: first in man

**DOI:** 10.1101/19000711

**Authors:** Marco Caversaccio, Wilhelm Wimmer, Juan Anso, Georgios Mantokoudis, Nicolas Gerber, Christoph Rathgeb, Daniel Schneider, Jan Hermann, Franca Wagner, Olivier Scheidegger, Markus Huth, Lukas Anschuetz, Martin Kompis, Tom Williamson, Brett Bell, Kate Gavaghan, Stefan Weber

**Author notes:** **Corresponding author:** Wilhelm Wimmer, PhD, Hearing Research Laboratory, ARTORG Center for Biomedical Engineering Research, Murtenstrasse 50, University of Bern, Bern, Switzerland.

## Abstract

To demonstrate the feasibility of robotic middle ear access in a clinical setting, nine adult patients with severe-to-profound hearing loss indicated for cochlear implantation were included in this clinical trial. A keyhole access tunnel to the tympanic cavity and targeting the round window was planned based on preoperatively acquired computed tomography image data and robotically drilled to the level of the facial recess. Intraoperative imaging was performed to confirm sufficient distance of the drilling trajectory to relevant anatomy. Robotic drilling continued toward the round window. The cochlear access was manually created by the surgeon. Electrode arrays were inserted through the keyhole tunnel under microscopic supervision via a tympanomeatal flap. All patients were successfully implanted with a cochlear implant. In 9 of 9 patients the robotic drilling was planned and performed to the level of the facial recess. In 3 patients, the procedure was reverted to a conventional approach for safety reasons. No change in facial nerve function compared to baseline measurements was observed. Robotic keyhole access for cochlear implantation is feasible. Further improvements to workflow complexity, duration of surgery, and usability including safety assessments are required to enable wider adoption of the procedure.

## INTRODUCTION

Advances in image guidance, robotic technology and minimally-invasive techniques offer an opportunity to transform inner ear surgery from open procedures to keyhole approaches. Over four decades after the description by House [1], conventional cochlear implant (CI) surgery remains essentially unchanged. The success story of CIs with about 600.000 implant users worldwide, shows the procedure is widely considered safe and effective [2-4]. Nevertheless, alternative implantation techniques to further improve patient outcomes such as reduced mastoidectomy under endoscopic supervision [5] and endaural implantation techniques have been proposed (e.g., the pericanal, suprameatal, or Veria approaches) [6-8]. However, mastoidectomy followed by a posterior tympanotomy remains the standard surgical approach to the inner ear for cochlear implantation.

Keyhole access CI surgery has become an active area of change for the procedure, with a view that image-guided, minimally-invasive and robotic equipment could be the starting point for a dedicated robotic CI procedure that could facilitate standardization of CI surgery and potentially impact hearing outcomes. Labadie et. al investigated a stereotactic frame-based keyhole intervention in eight CI patients [9]. Our group has developed the concept of robotic cochlear implantation (RCI) including elements of image-based procedural planning, robotic middle and inner ear access and finally robotic CI insertion, which aims to enable optimization and standardization of care. Following on from the first-in-man procedure [10], a clinical trial was carried out to test the hypothesis that a robotic and task-autonomous drill protocol can be applied to return the required geometric accuracy to enable a keyhole approach for cochlea implantation on varying anatomy. More specifically, elements such as the i) pre-operative planning process, the ii) multi-layer safety architecture and iii) tunnel based electrode insertion were clinically evaluated. Here we present the concluding report on the robotic middle ear access procedure carried out on a clinical pilot population of nine patients.

## METHODS

### Study Design

We performed a non-randomized single-center first-in-man trial to evaluate the clinical feasibility of RCI (Figure 1). The study protocol was approved by the local institutional review board (Ethics Commission of Bern, KEK-BE PB_2017-00312) and regulatory body (Swissmedic, Nr. 2013-MD-0042, EUDAMED CIV-13-12-011779) and registered (clinicaltrials.gov identifier: NCT02641795, trial registration on December 29^th^, 2015). Recruitment took place between 01.07.2016 and 22.08.2018 and surgeries took place between 14.07.2016 and 15.02.2018. All study procedures were performed at a tertiary referral hospital (Inselspital, Bern) in accordance with relevant guidelines and regulations. CI candidacy was evaluated according to a routine patient work-up, including medical, neuroradiological, and audiological assessment. Study-specific procedures consisted of screening, facial nerve function baseline testing at one day before surgery, the robotic intervention, computed tomography (CT) imaging one day after surgery, and follow-up testing 10 to 14 days after surgery. Clinical outcomes were assessed at 1 day, 2 weeks and 1 month after surgery. Safety of the trial was monitored by a board of three independent external reviewers.

**Figure 1.**
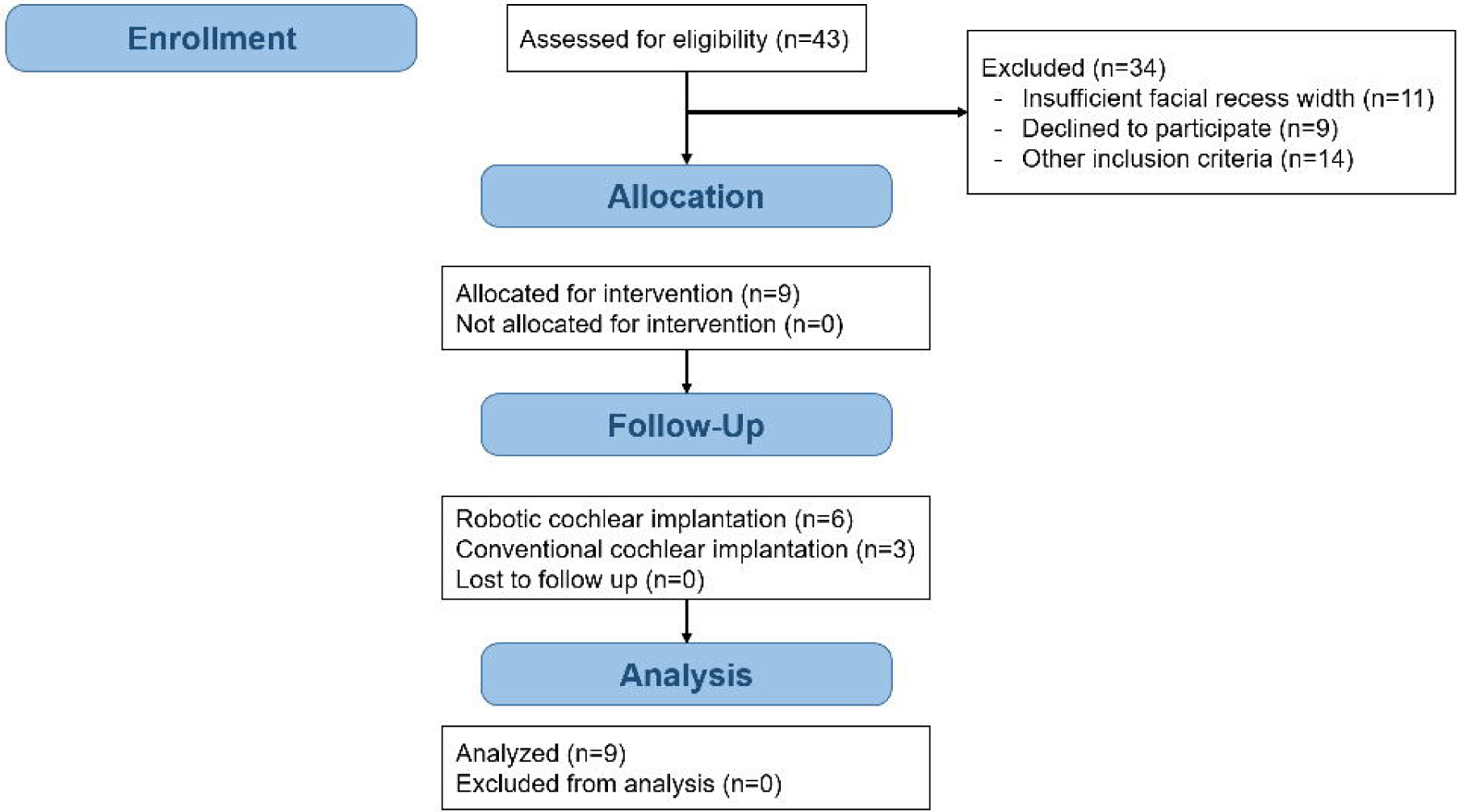
Flowchart for non-randomized trial design.

### Study Participants

Forty-three subjects were screened in our department as part of the CI candidacy assessment routine. Besides general accordance with conventional eligibility for CI surgery, subjects had to be 18 years or older, fluent in German or French, have a sufficient facial recess size (i.e., at least 2.5 mm, allowing for 0.4 mm safety margin to the facial nerve and 0.3 mm to the chorda tympani using a 1.8 mm drill bit). Exclusion criteria were: pregnancy, anatomical malformations of the middle or inner ear or abnormal course of the facial nerve. Existing preoperative CT datasets of the temporal bone were used to assess anatomical conditions and facial recess size [10]. In total, nine subjects gave written informed consented and were subsequently enrolled in the study (Table 1).

**Table 1.** Study participant details.

| ID | Age,<br>years | Gender | Side | Unaided PTA,<br>dB HL | Etiology | Hearing loss<br>duration, years | Electrode array |
| --- | --- | --- | --- | --- | --- | --- | --- |
| 01 | 51 | female | right | 120 | Cogan syndrome | 26 | Flex<sup>24</sup> |
| 02 | 49 | male | right | 94 | Morbus Menière | 22 | Flex<sup>28</sup> |
| 03* | 39 | female | left | 106 | Progressive hearing loss | 10 | Flex<sup>28</sup> |
| 04 | 68 | female | right | 71 | Progressive hearing loss | 12 | Flex<sup>28,CMD</sup> |
| 05 | 71 | female | right | 88 | Sudden deafness | 5 | Flex<sup>28</sup> |
| 06* | 59 | female | left | 89 | Progressive hearing loss | 7 | Flex<sup>28,CMD</sup> |
| 07 | 60 | male | right | 90 | Progressive hearing loss | 13 | Flex<sup>24</sup> |
| 08 | 73 | female | left | 86 | Morbus Menière | 26 | Flex<sup>28</sup> |
| 09 | 61 | male | right | 108 | Congenital | 20 | Flex<sup>28</sup> |

\*existing cochlear implant in contralateral ear.
PTA = pure tone average over 0.5/1/2/4 kHz; CMD = custom made device with shorter electrode lead.

### Baseline Testing

The motor portion of the facial nerve was evaluated using the standard Sunnybrook composite score [11]; and facial nerve conduction studies. The facial nerve was stimulated supramaximal at the mandibular angle, and amplitude and latency of the compound muscle action potentials were recorded using surface electrodes from the frontal, nasal, and mental muscles [12].

### Patient Preparation and intervention planning

The complete intervention was performed under general anesthesia. A 5 cm long retroauricular incision was created. A physical template was used to mark and insert four bone fiducial screws serving as artificial landmarks for patient-to-plan registration [13]. Patients were then transferred for preoperative CT imaging (Somatom CT, Siemens, Germany; voxel size: 0.156 × 0.156 × 0.2 mm^3^; 120 kVp) in the neuroradiological department to confirm correct insertion of the four implanted screws. Using a specific planning software [14], the team of the responsible surgeon, neuroradiologist and a trained computer engineer conducted next the procedural planning. First, the bony part of the external auditory canal, the facial nerve, the chorda tympani and the ossicles were annotated in the image data. Next, the access trajectory through the facial recess (diameter 1.8 mm distal and 2.5 mm proximal) from the mastoid surface to the center of the round window membrane, providing for an optimal insertion angle [15], was identified and defined in the image data. Meanwhile, the patient was transferred back to the operating room and prepared for the procedure [10]. The patient head was non-invasively constrained using a task specific headrest. Two sets of paired needle electrodes were inserted into the periorbital and perioral muscles to provide for facial nerve monitoring. To compensate for respiratory motion, the patients head was tracked via a skull attached dynamic reference base, aligned with the systems tracking camera [16].

### Robotic Middle Ear Access

Upon patient-to-image registration, a task specific robotic system [17,16] was used to drill the access tunnel (Figure 2) in 3 phases: (i) drilling from the surface of the mastoid bone until 3 mm before the facial nerve, (ii) passing through the facial recess, and (iii) completing the access to the middle ear cavity. Robotic drilling was performed in steps (with complete extraction of the drill bit between steps to allow for cooling and cleaning) with a feed forward rate of 0.5 mm per second and with 1000 revolutions per minute. Drilling increments of 0.5 mm and 2 mm were used for the passage of the facial recess and during the less critical phases drill phases i) and iii) respectively.

**Figure 2.**
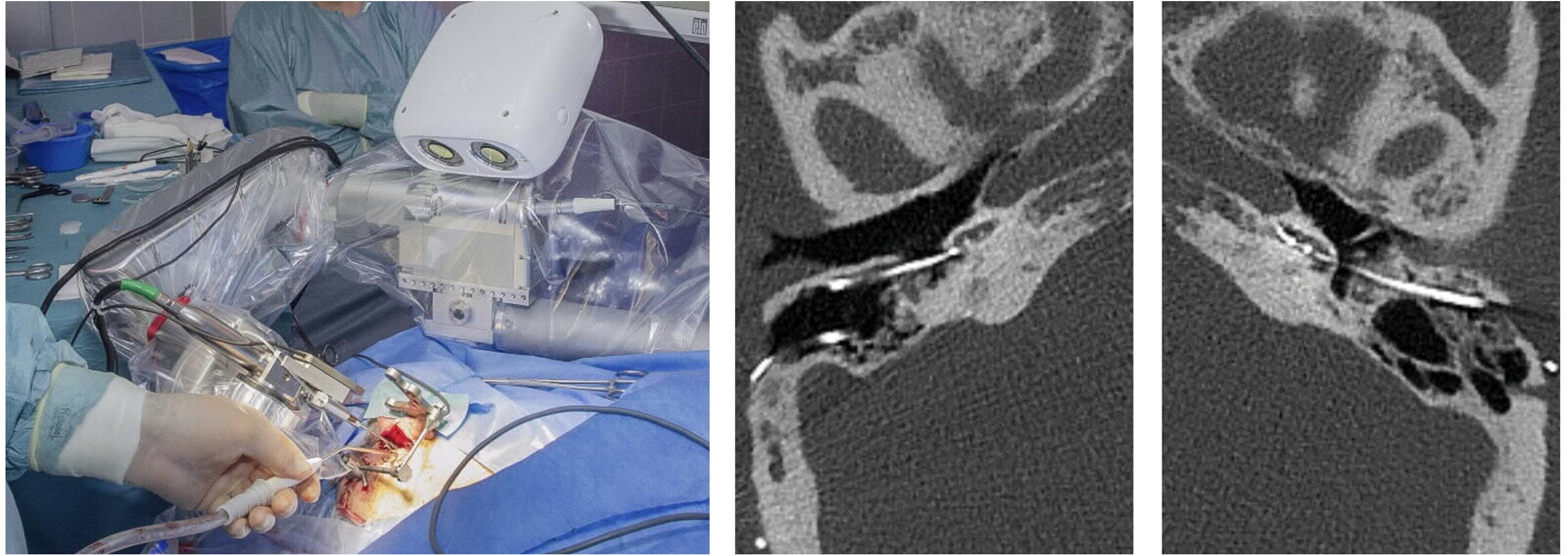
(left) The robotic system with patient. (right) Comparison between conventional and robotic procedure in postoperative computed-tomography slices (subject 06).

### Intraoperative imaging

Upon reaching the level of the facial nerve, the robot was moved away from the operating table and a titanium rod was inserted into the drilled tunnel. At that point the temporal bone region was repeated imaged with a low-dose radiation, cone-based CT scanner for use in operating rooms (xCAT, Xoran, Ann Arbor, USA; voxel resolution: 0.3 × 0.3 × 0.3 mm^3^, 120 kVp, 6 mA; Figure 3 left). The acquired image data were used to measure the distances of the drill tunnel to surrounding anatomical structures by using an automatic detection algorithm [18]. A safe trajectory was also confirmed manually on the image data by a present neuroradiologist. The distance of the drill tunnel to relevant anatomy was also assessed by the systems integrated force-density pose estimation calculation [16,19]. Subsequently, the responsible surgeon decided whether the robotic procedure would continue (i.e., phase ii) or be reverted to a conventional approach.

**Figure 3.**
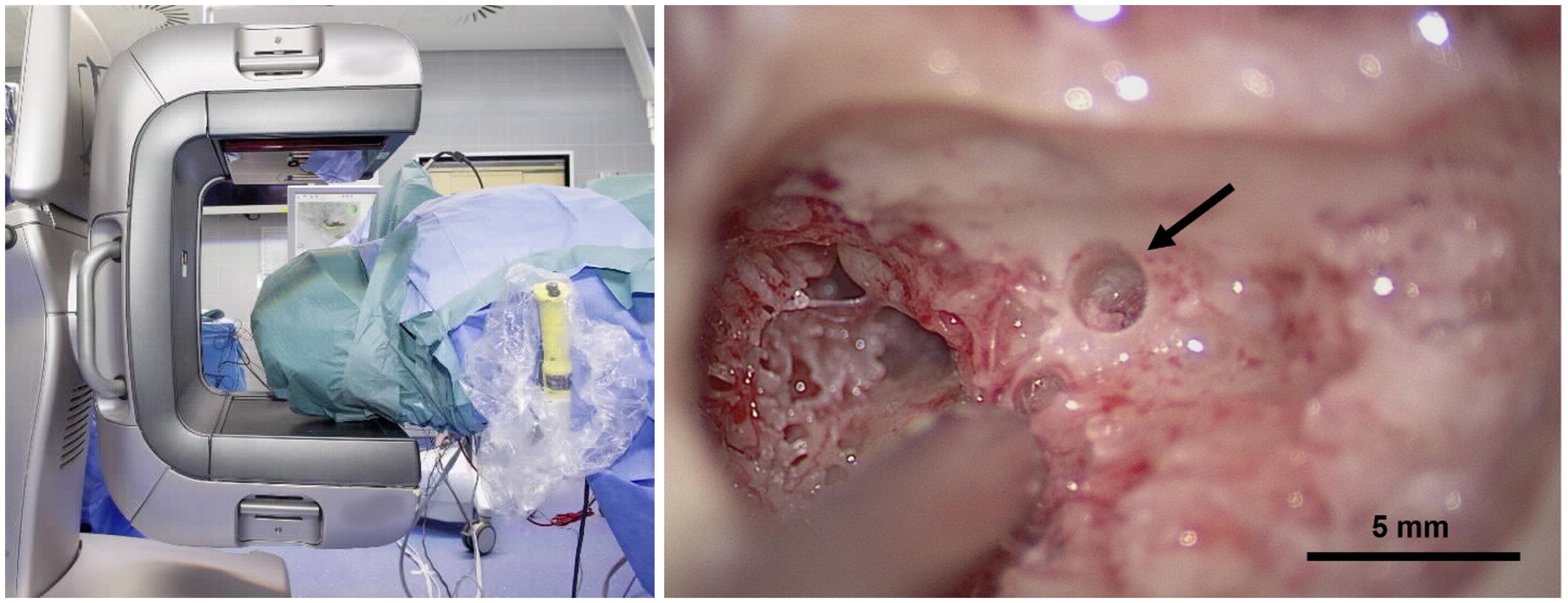
(left) Patient prepared for intraoperative CBCT imaging. (right) Microscopic inspection of the robotically drilled tunnel (arrow) after reversion to conventional procedure including a mastoidectomy (subject 02).

### Electromyography for facial nerve monitoring

Integrity of the facial nerve was monitored continuously through i) measuring potential electromyography (EMG) discharges elicited by mechanical irritation of the facial nerve during the complete drilling and ii) analyzing compound muscle action potentials from multipolar stimulation specifically when passing the facial nerve [12,20].

### Implantation

After completion of the access tunnel, the landmark screws and the dynamic reference base were removed. The tunnel was cleaned using irrigation and suction. Tunnel alignment with the round window was inspected with a sialendoscope (Karl Storz, Tuttlingen, Germany). The retroauricular incision was extended inferiorly and a tympanomeatal flap was created as an auxiliary access to the tympanic cavity [21]. The round window niche was microscopically visualized through the external auditory canal. Through the tympanomeatal flap, the bony overhang of the round window was removed using a skeeter drill (Bien-Air, Biel, Switzerland). Next, the implant bed was prepared. As opposed to conventional CI surgery, the excess lead of the implant cannot be accommodated within the mastoidectomy with the minimally invasive approach. Therefore, a superficial well (2 mm in depth) was milled into the cortex of the bone to enable safe embedding. This step was not required to the same extent with the custom-made device implants featuring shortened leads (subjects 04 and 06). The middle ear cavity and the implant bed were cleaned to avoid intrusion of bone dust and blood into the cochlea during electrode array insertion. The round window membrane was opened with a micro-needle. If required, a custom-made insertion guide tube, was placed inside the tunnel to assist insertion [22] and provide against unwanted contamination of the electrode with blood. Sodium hyaluronate was applied as lubricant (ProVisc^®^, Alcon, Rotkreuz, Switzerland). The electrode array was slowly inserted until the first point of resistance. After insertion, the guide tube was removed and the round window niche was sealed with fatty tissue. The excess lead was fixed at the top of the tunnel using bone wax (Ethicon, Somerville, US). Following implant telemetry, the wound was closed. Implant body management was adapted from the conventional CI procedure.

### Outcome measures

Drilling accuracy (primary outcome) was measured in available intraoperative CB-CT scans at the level of the facial recess as the absolute centerline displacement of the planned versus the drilled tunnel. The number of fully completed robotic accesses to the tympanic cavity, the total procedural durations and durations of all sub-procedures were recorded. Postoperative high-resolution CT scans of the temporal bone were used to measure the angular insertion depth (in degrees) and the implanted scala (secondary outcomes). The surgical follow up included assessment of pain (visual analog scale) and any potential clinical complications. Functional preservation of the facial nerve was postoperatively assessed relative to the preoperative baseline measurements. Structural preservation of the chorda tympani was confirmed by a neuroradiologist in the standard postoperative high-resolution CT scan of the temporal bone with similar scanning parameters to the preoperative CT. Audiological fitting was performed according to our standard routine, i.e., activation and initial fitting at 1 month and consecutive fitting sessions at up to 12 months after the surgery. Audiological evaluation included the number of activated channels, aided sound field thresholds (pure tone average over 0.5/1/2/4 kHz, in dB HL), aided word recognition scores (in %) for monosyllables (at 65 dB SPL) and numbers (at 70 dB SPL). If applicable, the degree of hearing preservation (in %) was quantified [23].

## RESULTS

### Feasibility

Of 43 initially assessed subjects planned for cochlear implantation, 29 patients were screened for facial recess size, the other 14 patients were excluded because one or more of the other inclusion criteria were unmet. Eighteen of the 29 patients had a sufficient facial recess size and were eligible for the study. Of those, nine patients consented to participation in the trial. In all nine patients, a safe access tunnel to the level of the facial recess was planned and drilled. The complete robotic procedure including drilling through the facial recess was performed in 6 of 9 patients. Insufficient distances to the facial nerve (< 0.3 mm) and the tympanic membrane (< 0.1 mm) were detected in the available intraoperative image data in patients 8 and 9 respectively. Hence, procedures were reverted to a conventional transmastoid posterior tympanotomy. In subject 2, the patient’s mastoid region could not be imaged due to workspace limitations and a compressed cervical spine region. Hence, as intraoperative image data was required for confirmation of sufficient tool clearance by study design, this case was also reverted to the standard procedure. In all 3 reverted cases clearance of the drill trajectory to the facial nerve was confirmed microscopically during mastoidectomy (Figure 3 right). Drilling accuracy, measured as the deviation between the planned and the drilled tunnel at the level of the facial recess, was 0.21 mm ± 0.09 mm (Table 2) which is in-line with preclinical validation [17]. All nine patients were implanted with a CI (SYNCHRONY, MED-EL, Innsbruck, Austria) under full preservation of facial nerve function. No abnormal EMG activity or low stimulation thresholds were identified during the entire robotic drilling phases. Implanted subjects neither showed a change in Sunnybrook composite score nor in facial nerve conduction study parameters compared to baseline measurements. In all cases, preservation of the chorda tympani was confirmed in the postoperative image evaluation. Overall, procedural blood loss was less than 50 ml loss in eight of nine cases, and 170 ml in one case. One day after surgery, eight of nine patients reported pain levels below or equal 2 using a visual analog scale and were painless in the follow-up examinations. In the six patients with a complete robotic middle ear access, the electrode array was inserted through the drilled tunnel with an angular insertion depth of 501° ± 94° (Table 2). In subject 1, plaque formation in the cochlear basal turn (Cogan syndrome) prevented insertion into the scala tympani and resulted in scala vestibuli placement as predicted in the preoperative planning [10]. Scala tympani insertion was reported in all other subjects. All patients were discharged from hospital one day after surgery.

**Table 2.**
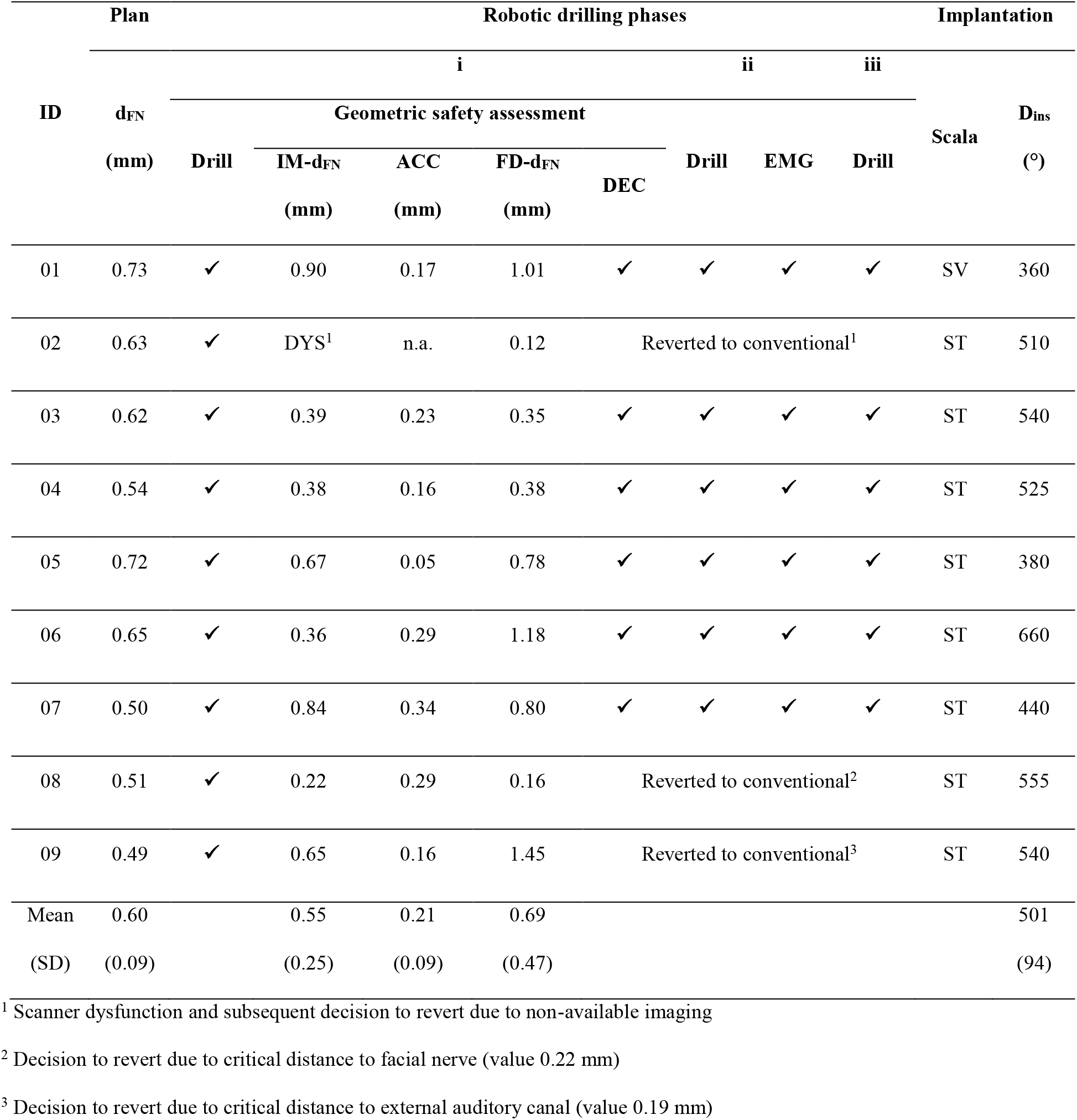
Summary of results. IM-d_FN_ is the distance tunnel to facial nerve based on the intraoperative imaging; ACC = effective drilling accuracy at the level of the facial recess; FD-d_FN_ = estimated distance of the drill tunnel to the facial nerve using force-density correlation; DEC = Confirmation for sufficient geometric clearance; D_ins_ = angular insertion depth; SV = scala vestibuli; ST = scala tympani.

### Duration

The total average procedure duration (skin to skin) was 4:05 hours (min/max. 3:15/5:00 hours). The averaged sub-procedural times were: screw insertion (13 min), patient transfer and preoperative CT imaging (29 min), surgical planning (37 min), patient preparation (60 min, performed simultaneously with surgical planning), patient registration (8 min), robotic drilling to the level of the facial recess (phase (i), 6 min), intraoperative imaging and analysis (54 min), robotic drilling through the facial recess with intermittent facial nerve stimulation and monitoring (phase (ii), 16 min), drilling to the middle ear cavity (phase (iii) (5 min), tympanomeatal flap (17 min), implant bed preparation (9 min), cochlear access (14 min), CI electrode array insertion (6 min), and implant fixation and wound closure (8 min).

### Audiological Outcome

Aided sound field hearing thresholds as well as aided word recognition for monosyllables and numbers showed clear benefit after CI activation (Table 3). The average word recognition scores for numbers were 54% (N=9), 69% (N=9), 66% (N=5), and 72% (N=5), at 1, 3, 6 and 12 months after surgery, respectively. The patients achieved monosyllabic word recognition scores of 23% (N=9), 39% (N=9), 50% (N=5), and 56% (N=5), respectively. In the patients with low frequency residual hearing, minimal to partial hearing preservation after surgery was achieved (Table 3).

**Table 3.**
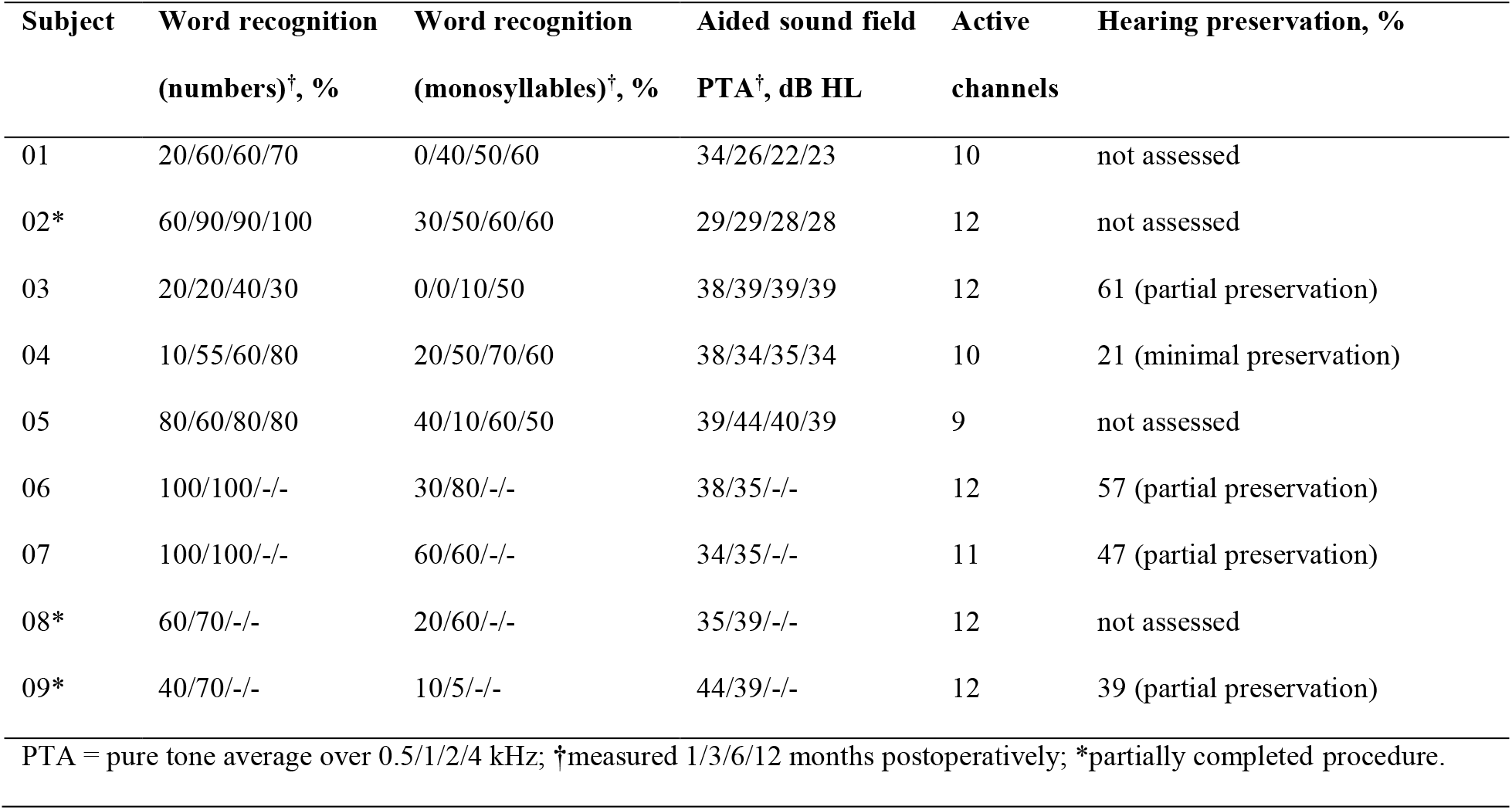
Audiological outcomes.

| Subject | Word recognition<br>(numbers)<sup>†</sup>, % | Word recognition<br>(monosyllables)<sup>†</sup>, % | Aided sound field<br>PTA<sup>†</sup>, dB HL | Active<br>channels | Hearing preservation, % |
| --- | --- | --- | --- | --- | --- |
| 01 | 20/60/60/70 | 0/40/50/60 | 34/26/22/23 | 10 | not assessed |
| 02* | 60/90/90/100 | 30/50/60/60 | 29/29/28/28 | 12 | not assessed |
| 03 | 20/20/40/30 | 0/0/10/50 | 38/39/39/39 | 12 | 61 (partial preservation) |
| 04 | 10/55/60/80 | 20/50/70/60 | 38/34/35/34 | 10 | 21 (minimal preservation) |
| 05 | 80/60/80/80 | 40/10/60/50 | 39/44/40/39 | 9 | not assessed |
| 06 | 100/100/-/- | 30/80/-/- | 38/35/-/- | 12 | 57 (partial preservation) |
| 07 | 100/100/-/- | 60/60/-/- | 34/35/-/- | 11 | 47 (partial preservation) |
| 08* | 60/70/-/- | 20/60/-/- | 35/39/-/- | 12 | not assessed |
| 09* | 40/70/-/- | 10/5/-/- | 44/39/-/- | 12 | 39 (partial preservation) |

PTA = pure tone average over 0.5/1/2/4 kHz; <sup>†</sup>measured 1/3/6/12 months postoperatively; \*partially completed procedure.

## DISCUSSION

Based on the criterion of sufficient width of the facial recess, 62% of screened patients (18 of 29) were eligible. This compares favorably with a previous estimate of 47% [24]. The feasibility of the surgeon driven, task-autonomous robotic drilling procedure was demonstrated in 6 of 9 patients. In 3 patients, the RCI workflow decision-making led the team to revert the procedure to a conventional CI. The study shows that micro-surgical robotic technology can be deployed in a clinically resilient manner and across varying patient anatomies to deliver geometrically-accurate keyhole, access to the inner ear. In subject 02, the workspace limitation of the intraoperative CB-CT scanner caused reversion to a conventional procedure, which can be avoided by employing pre-surgical anatomy assessments or alternative imaging technologies with greater imaging volumes. In subjects 08 and 09, the deviated drill tunnel (Table 2) led to critical proximity to the facial nerve and auditory canal wall and thus the procedures were reverted. Although these two subjects did not have a full, robotic drill path past the facial nerve, the procedures demonstrated the effectiveness of intraoperative imaging at the decision point 3 mm before being level with the facial nerve, as a key safety feature. The previously identified and validated drill geometry configuration and drill process parametrization [25] demonstrated feasibility and safety in all 9 subjects. In the six cases with a complete robotic middle ear access, the force density drill pose safety assessment corroborated the safe passage determined on intraoperative images. EMG based distance measurements made intermittently and while passing the facial recess were always conclusive with the imaging-based tool-to-nerve distance assessments and resulted in safe continuation of the robotic drilling procedure [12]. In all implanted patients, both the clinical and electrophysiological facial nerve function remained intact compared to baseline measurements. Time required for intraoperative imaging confirmation of sufficient instrument-to-nerve distance was 54 mins. Nonetheless, workflow optimization and complete intraoperative integration of imaging and image analysis technology (both for planning and confirmation) into the workflow and routine OR work will lead to significant time and cost reductions. During this trial an auxiliary access was required for three reasons: (i) drilling of the cochlear access, in our case by removal of the bony overhang, (ii) as backup access in case of bleeding or unanticipated problems during array insertion, and (iii) sealing of the cochlear access after insertion completion. To facilitate the lifting of the tympanomeatal flap, an L-shaped retroauricular incision was replaced by a C-shaped incision after the first case. In future, optimized electrode array designs and robotically performed inner ear access and electrode array insertion may remove the need for this secondary access. Although, not being an endpoint of this trial, we demonstrated that hearing preservation can be achieved with the RCI procedure. Audiological outcomes compare favorably to conventional cochlear implantation [26,27], however further study is required.

The presented work introduces task-autonomous surgical robotics to the field of otological microsurgery (autonomy level 2) [28]. Robotic technology offers possibilities to overcome human operator limitations to provide for reproducible, minimally invasive cochlear access and ultimately a deliberate and accurate electrode insertion process, potentially widening CI patient eligibility in the future. We consider the work presented as a first step towards this goal and believe to have demonstrated feasibility of the overall approach in a sufficient variety of patient anatomies and workflow iterations. Interestingly, a robotic keyhole access renders direct visual supervision of the actual drilling process impossible. Hence, safety elements such as EMG-based facial nerve monitoring and intraoperative imaging were utilized to confirm correct drill alignment. To ensure safety of the robotic access and to demonstrate the efficacy of the applied safety measures, independent clinical trials with larger patient numbers need to be performed. Compared to conventional cochlear implantation, the presented approach is more time-consuming and labor-intensive. As with all novel surgical techniques, an increased average duration of the surgery owing to learning curve and the execution of safety procedures is to be expected throughout the first cases. Most prominently and because of the underlying technological complexity, every step in the workflow was carefully co-checked, monitored and confirmed by the multi-disciplinary team, resulting in a reduced overall workflow efficiency. In addition, preoperative high-resolution CT scans were conducted outside the OR (resulting in patient preparation and transportation) prolonging the overall procedure time. Additionally, time was required for intraoperative imaging together with the necessary image data transfer, peer assessment and subsequent decision making. Further integration of intraoperative imaging devices will drastically reduce the time needed for pre- and intraoperative imaging and image processing. To introduce a complete robotic cochlear implantation approach, we are currently developing and investigating solutions for robotic inner ear access, robotic electrode insertion, multi-port scenarios [29], narrower drills (i.e. 1mm to 1.4 mm) with integrated monitoring electrodes and ultimately robotics compliant CI implant technology.

## Data Availability

All relevant data are within the manuscript and its Supporting Information files.

## ACKNOWLEDGMENTS

The authors thank Dr. Simona Negoias, Dr. Cilgia Dür, and Gianni Pauciello, Department of ENT, Head and Neck Surgery, and Dr. Stefan Henle, Department of Anesthesiology and Pain Medicine, Inselspital, Bern University Hospital, University of Bern, Bern, Switzerland for their contributions to the study. The authors acknowledge financial and in-kind contributions made possible by MED-EL GmbH (Innsbruck Austria), CAScination AG (Bern Switzerland) and Xoran Technologies (Ann Arbor, USA).

